# Amyloid-β CSF/PET discordance vs tau load 5 years later: It takes two to tangle

**DOI:** 10.1101/2020.01.29.20019539

**Authors:** Juhan Reimand, Lyduine Collij, Philip Scheltens, Femke Bouwman, Rik Ossenkoppele, for the Alzheimer’s Disease Neuroimaging Initiative

**Affiliations:** Department of Neurology & Alzheimer Center Amsterdam, Amsterdam Neuroscience, Vrije Universiteit Amsterdam, Amsterdam UMC, Amsterdam, the Netherlands; Department of Health Technologies, Tallinn University of Technology, Tallinn, Estonia; Radiology Centre, North Estonia Medical Centre, Tallinn, Estonia; Department of Radiology and Nuclear Medicine, Amsterdam Neuroscience, Vrije Universiteit Amsterdam, Amsterdam UMC, Amsterdam, the Netherlands; Clinical Memory Research Unit, Lund University, Lund, Sweden

**Author notes:** Corresponding Author: Rik Ossenkoppele. Data used in preparation of this article were obtained from the Alzheimer’s Disease Neuroimaging Initiative (ADNI) database (adni.loni.usc.edu). As such, the investigators within the ADNI contributed to the design and implementation of ADNI and/or provided data but did not participate in analysis or writing of this report. A complete listing of ADNI investigators can be found at: http://adni.loni.usc.edu/wp-content/uploads/how_to_apply/ADNI_Acknowledgement_List.pdf.

## Abstract

**OBJECTIVE:** To investigate the association between discordant amyloid-β PET and CSF biomarkers at baseline and the emergence of tau pathology 5 years later.

**METHODS:** We included 730 ADNI participants without dementia (282 cognitively normal, 448 mild cognitive impairment) with baseline [^18^F]Florbetapir PET and CSF Aβ42 available. Amyloid-β CSF/PET status was determined at baseline using established cut-offs. Longitudinal data was available for [^18^F]florbetapir (Aβ) PET (baseline to 4.3±1.9 years), CSF (p)tau (baseline to 2.0±0.1 years), cognition (baseline to 4.3±2.0 years), and [^18^F]flortaucipir (tau) PET (measured 5.2±1.2 years after baseline to 1.6±0.7 years later). We used linear mixed modelling to study the association between amyloid-β CSF/PET status and tau pathology measured in CSF or using PET. Additionally, we calculated the proportion of CSF+/PET-participants who during follow-up (i) progressed to amyloid-β CSF+/PET+, and/or (ii) became tau-positive based on [^18^F]flortaucipir PET.

**RESULTS:** Amyloid-β CSF+/PET+ (N=318) participants had elevated CSF (p)tau levels and worse cognitive performance at baseline, while CSF+/PET-(N=80) participants were overall similar to the CSF-/PET-(N=306) group. Five years after baseline, [^18^F]flortaucipir PET uptake in the CSF+/PET-group (1.20±0.13) did not differ from CSF-/PET-(1.18±0.08, p=0.69), but was substantially lower than CSF+/PET+ (1.48±0.44, p<0.001). Of the CSF+/PET-subjects, 21/64 (33%) progressed to amyloid-β CSF+/PET+, whereas only one (3%, difference p<0.05) became tau-positive based on [^18^F]flortaucipir PET.

**CONCLUSIONS:** Sufficient amyloid-β load detectable by both CSF and PET is required before substantial tau deposition emerges. Compared to participants with abnormal amyloid-β levels on PET and CSF, the CSF+/PET-group has a distinctly better prognosis.

## Introduction

Amyloid-β plaques and neurofibrillary tau tangles are considered the pathological hallmarks of Alzheimer’s disease (AD).^1^ Amyloid-β pathology can be measured *in vivo* directly by quantifying the fibrillary depositions using positron emission tomography (PET), or indirectly by detecting the decrease of soluble Aβ42 in cerebrospinal fluid (CSF). Although these two measures are sometimes considered interchangeable,^2–4^ 10-20% cases show discordant results, especially at earlier stages of AD.^5–7^ Therefore, it has been proposed that decreased Aβ42 in CSF without significant tracer uptake on PET (i.e. amyloid-β CSF+/PET-) marks the pathological beginnings of amyloid-β accumulation.^8,9^ This provides a powerful model to study the dynamic changes in amyloid-β as well as tau pathology during the earliest stages in the disease course of AD. Although CSF tau biomarkers have been available for over two decades,^10^ tau PET using radiotracers such as [^18^F]flortaucipir^11,12^ has only more recently been developed. Tau PET offers the unique opportunity to study the spatial distribution of tau aggregates *in vivo*.

An open question to date is whether isolated amyloid-β positivity in CSF is followed by significant tau deposition already at this stage, or whether it will be subsequent to more advanced amyloid-β pathology detectable by both modalities (i.e. CSF and PET). As tau has a stronger correlation to neurodegeneration and cognitive function than amyloid-β accumulation,^13,14^ this is of high clinical relevance. Furthermore, a better understanding of the interplay between the AD hallmark pathologies in early disease stages is crucial for the timing of interventions, as emerging treatments will likely be most effective when substantial neurodegeneration has not yet developed.^15^

## Methods

### Participants

Data for this study was downloaded from the Alzheimer’s Disease Neuroimaging Initiative (ADNI) website (http://adni.loni.usc.edu/), which also includes information about ADNI inclusion criteria and the procedure of biomarker acquisition.^16^ We selected all ADNI participants, who had at least one [^18^F]florbetapir amyloid-β PET scan and a CSF Aβ42 analysis available within one year. The diagnosis closest to baseline [^18^F]florbetapir PET within one year was used as the baseline diagnosis. In total, we included 730 pre-dementia subjects, of whom 282 were cognitively normal (CN) and 448 had mild cognitive impairment (MCI) at baseline.

We used Mini-Mental State Examination (MMSE) to assess global cognition and composite z-scores to assess memory^17^ and executive functioning.^18^ Longitudinal cognitive assessment was available for 711 (97%) subjects, with a median follow-up time of 4.2 [interquartile range (IQR): 2.9, 5.9] years. Similarly, a follow-up diagnosis was available for 724 (99%) participants with a median interval between baseline and follow-up of 4.1 [IQR: 2.2, 5.9] years.

### Standard protocol approvals, registrations, and patient consents

Written informed consent was obtained for all participants, and study procedures were approved by the institutional re-view board at each of the participating centers. ADNI is listed in the ClinicalTrials.gov registry (ADNI-1: NCT00106899; ADNI-GO: NCT01078636; ADNI-2: NCT0123197).

### [^18^F]florbetapir positron emission tomography

Acquisition and processing of [^18^F]Florbetapir PET using Freesurfer was performed as described previously.^19,20^ At least one follow-up PET scan was available for 579 (79%) of participants with a median follow-up time of 4.1 [IQR: 2.1, 5.9] years. We used a neocortical composite score provided by ADNI consisting of the mean uptake in the frontal, cingulate, parietal and temporal regions. We created standardized uptake value ratios (SUVR) using whole cerebellum as the reference region and used an SUVR cut-off of 1.11 to determine binarized amyloid-β status based on PET.^21,22^ For longitudinal linear mixed modelling, we additionally used SUVR values in 34 regions from the Desikan-Killiany atlas using (i) the whole cerebellum,^23^ and (ii) a composite white matter region as reference regions, because the latter has been shown to be more reliable in longitudinal analyses.^24^ Finally, we used an early composite region identified in a recent study^25^ (including bilateral precuneus, posterior cingulate, insula, and medial and lateral orbitofrontal regions) to capture the early accumulation of amyloid-β.

### Cerebrospinal fluid

Lumbar punctures were performed as previously described.^26^ CSF samples were frozen on dry ice after collection and transported to the UPenn Medical Center ADNI Biomarker Core laboratory. Thereafter, 0.5mL aliquots were prepared and stored in polypropylene tubes at - 80°C. CSF samples were analyzed for Aβ42, total tau (t-tau) and phosphorylated tau (p-tau) using the AlzBio3 assays (Fujirebio) on the xMAP platform (Luminex). In case samples were reanalyzed, we used the median value of those results. A cutoff of 192 pg/mL was used to determine amyloid-β status based on CSF.^26,27^ Longitudinal CSF analyses were available for 297 (41%) of participants with the median follow-up time of 2.0 [IQR: 2.0, 2.0] years.

### [^18^F]flortaucipir positron emission tomography

[^18^F]Flortaucipir PET was performed at each ADNI site according to standardized protocols. Images were acquired via 4×5 minute frames from 80 to 100 minutes after injecting ∼370 MBq of [^18^F]flortaucipir. [^18^F]flortaucipir PET was available for 253 (35%) participants and was performed a median of 5.2 [IQR: 4.2, 6.1] years after baseline [^18^F]florbetapir PET, allowing measuring tau pathology at a significantly later time-point. Of these participants, 110 had one follow-up scan after 1.3 [IQR: 1.0, 2.1] years. We used Freesurfer-defined Desikan-Killiany atlas regions provided by ADNI that were created by co-registering the [^18^F]flortaucipir image with a previously parcellated and segmented magnetic resonance imaging MPRAGE from the same time.^28^ Thereafter, we created three bilaterally volume-weighted composite regions to cover the full spectrum of tau aggregation: entorhinal cortex, temporal meta-ROI reflecting Braak stage I to IV (including entorhinal, parahippocampal cortex, amygdala, fusiform, inferior and middle temporal cortices), and Braak stage V and VI (including wider neocortical areas).^29–31^ Cut-offs (1.39, 1.34, and 1.28 SUVR, respectively) obtained using a similar PET pipeline were used to determine [^18^F]flortaucipir positivity.^32^

### Statistical analysis

We selected the first available [^18^F]florbetapir PET as baseline amyloid-β PET, and CSF Aβ42 closest in time within one year to the [^18^F]florbetapir PET as baseline CSF. Thereafter, we created four groups based on the binarized amyloid-β status on PET and CSF: concordantly amyloid-negative (CSF-/PET-), concordantly amyloid-positive (CSF+/PET+), discordantly amyloid-positive based on CSF (CSF+/PET-) or PET (CSF-/PET+). Participant groups were compared using Chi-squared tests, two samples t-tests and Wilcoxon Rank-Sum tests. Statistical analysis was performed using R software version 3.5.3.^33–36^

We used linear mixed modelling to investigate longitudinal changes for (i) regional amyloid-β burden assessed by [^18^F]florbetapir PET, (ii) tau pathology assessed by CSF t-tau, p-tau (measured from baseline) and [^18^F]flortaucipir PET (measured 5 years after baseline), and (iii) cognitive measures (MMSE, and ADNI memory and executive composite scores). The models included time in years as a continuous variable, amyloid-β CSF/PET group, and an interaction between time*CSF/PET group. All models also included terms for age and sex. The models predicting regional amyloid-β PET and tau pathology based on CSF or PET were additionally adjusted for MMSE to account for clinical disease severity. The models predicting cognitive test results were additionally adjusted for education. We used a random intercept and a random slope for all models. We first selected CSF-/PET-as the reference group and interpreted the main effect of CSF/PET group status (CSF-/PET+ and CSF+/PET-) in the models as difference at baseline, and the CSF/PET group*time interaction term as the change over time. Thereafter, we repeated this analysis with CSF+/PET+ as the reference group. We then performed Kaplan-Meier survival analyses to investigate the association between amyloid-β CSF/PET status and clinical progression by using both overall progression (CN to MCI or dementia, or MCI to dementia) and progression to dementia (CN or MCI to dementia) as events. We additionally used Cox Regression analyses to obtain *post hoc* Hazard Ratios for each of the amyloid-β CSF/PET profiles.

Finally, we performed two analyses in CSF+/PET-participants only. First, to investigate subthreshold levels of tau pathology in the amyloid-β CSF+/PET-group, we performed linear regression models with amyloid-β pathology measured by [^18^F]florbetapir PET (globally as well as in early accumulating regions^25^) at baseline as the outcome, and cross-sectional tau pathology measured by either CSF t-tau or p-tau at baseline, or the three composite regions of [^18^F]flortaucipir PET five years later as predictors. Secondly, we investigated our hypothesis that progression from amyloid-β CSF+/PET-to CSF+/PET+ occurs at a higher rate than progression to tau-positivity based on [^18^F]flortaucipir PET. We calculated the proportions of CSF+/PET-participants, who (i) during the follow-up period converted into the CSF+/PET+ group based on the last available [^18^F]florbetapir PET scan and CSF analysis, and (ii) whose last available [^18^F]flortaucipir PET was positive in any of the 3 composite regions. We compared these outcomes by testing for overlapping 95% confidence intervals on estimated proportions. Last available [^18^F]flortaucipir PET was performed a median of 6.0 [IQR: 5.5, 6.8] years after baseline and 0.0 [-2.0, 1.4] years from last available [^18^F]florbetapir PET.

### Data availability

All imaging, demographics, and neuropsychological data used in this article are publicly available and were downloaded from the ADNI website (www.adni.loni.usc.edu). Upon request, we will provide a list of ADNI participant identifications for replication purposes.

## Results

### Study participants

Of the study participants, 306 (42%) were CSF-/PET-, 80 (11%) CSF+/PET-, 26 (4%) CSF-/PET+ and 318 (44%) CSF+/PET+ **(Table 1)**. Characteristics were overall similar between CSF-/PET- and the two discordant groups. Participants in the CSF+/PET+ group had a higher proportion of *APOE* ε4 carriers, were more often diagnosed with MCI, had lower cognitive scores, higher CSF (p)tau levels at baseline and higher [^18^F]flortaucipir PET uptake five years later.

**Table 1.** Study participants

|  | CSF-/PET- | CSF+/PET- | CSF-/PET+ | CSF+/PET+ |
| --- | --- | --- | --- | --- |
| **n (%)** | 306 (42) | 80 (11) | 26 (4) | 318 (44) |
| **Sex, male (%)** | 161 (53)<sup>C</sup> | 48 (60)<sup>C</sup> | 6 (23)<sup>ABD</sup> | 165 (52)<sup>C</sup> |
| **Age (mean (SD))** | 72 (7)<sup>D</sup> | 72 (8) | 71 (6)<sup>D</sup> | 74 (7)<sup>AC</sup> |
| **Education (median [IQR])** | 17 [14, 18] | 16 [16, 18] | 16 [14, 18] | 16 [14, 18] |
| **APOE ε4 carriership (%)** | 53 (17)<sup>BD</sup> | 23 (32)<sup>AD</sup> | 8 (31)<sup>D</sup> | 159 (58)<sup>ABC</sup> |
| **Diagnosis, MCI (%)** | 155 (51)<sup>D</sup> | 41 (51)<sup>D</sup> | 13 (50)<sup>D</sup> | 239 (75)<sup>ABC</sup> |
| **[<sup>18</sup>F]florbetapir PET composite SUVR (median [IQR])** | 1.01 [0.98, 1.04]<sup>BCD</sup> | 1.05 [1.00, 1.08]<sup>ACD</sup> | 1.15 [1.13, 1.19]<sup>ABD</sup> | 1.36 [1.26, 1.49]<sup>ABC</sup> |
| **CSF Aβ<sub>42</sub> (median [IQR])** | 232 [213, 249]<sup>BCD</sup> | 164 [149, 181]<sup>ACD</sup> | 214 [204, 243]<sup>ABD</sup> | 136 [121, 154]<sup>ABC</sup> |
| **<u>Cognitive test scores (baseline):</u>** |  |  |  |  |
| **MMSE (mean (SD))** | 28.8 (1.4)<sup>D</sup> | 28.8 (1.4)<sup>D</sup> | 28.6 (1.6) | 28 (1.8)<sup>AB</sup> |
| **ADNI memory composite (mean (SD))** | 0.90 (0.68)<sup>D</sup> | 0.76 (0.64)<sup>D</sup> | 1.09 (0.48)<sup>D</sup> | 0.30 (0.71)<sup>ABC</sup> |
| **ADNI executive composite (mean (SD))** | 0.85 (0.85)<sup>BD</sup> | 0.55 (0.74)<sup>ACD</sup> | 1.13 (0.77)<sup>BD</sup> | 0.27 (0.86)<sup>ABC</sup> |
| **<u>CSF tau measures (baseline):</u>** |  |  |  |  |
| **CSF t-tau (median [IQR])** | 53 [42, 68]<sup>D</sup> | 55 [39, 76]<sup>D</sup> | 57 [48, 74]<sup>D</sup> | 93 [68, 135]<sup>ABC</sup> |
| **CSF p-tau (median [IQR])** | 25 [20, 35]<sup>D</sup> | 24 [19, 37]<sup>D</sup> | 26 [21, 43]<sup>D</sup> | 47 [35, 65]<sup>ABC</sup> |
| **<u>[<sup>18</sup>F]flortaucipir PET (5 years later):</u>** |  |  |  |  |
| **Entorhinal SUVR (median [IQR])** | 1.12 [1.06, 1.17]<sup>D</sup> | 1.13 [1.08, 1.19]<sup>D</sup> | 1.11 [1.06, 1.15]<sup>D</sup> | 1.40 [1.21, 1.61]<sup>ABC</sup> |
| **Temporal meta-ROI SUVR (median [IQR])** | 1.18 [1.13, 1.23]<sup>D</sup> | 1.18 [1.15, 1.23]<sup>D</sup> | 1.18 [1.16, 1.24]<sup>D</sup> | 1.36 [1.22, 1.58]<sup>ABC</sup> |
| **BRAAK V and VI SUVR (median [IQR])** | 1.06 [1.02, 1.10]<sup>CD</sup> | 1.08 [1.01, 1.10]<sup>D</sup> | 1.11 [1.07, 1.17]<sup>A</sup> | 1.16 [1.06, 1.25]<sup>AB</sup> |

A,B,C,D indicate differences ( $p < 0.05$ ) from other groups: A – difference from CSF-/PET-; B – difference from CSF+/PET-; C – difference from CSF-/PET+; D – difference from CSF+/PET+. Baseline diagnosis was either cognitively normal or mild cognitive impairment (MCI). Cognitive test scores were compared while adjusting for age, sex and education. False discovery rate was used to correct for multiple comparison. SUVR - standardized uptake value ratios.

### Accumulation of amyloid-β

First, we assessed regional [^18^F]florbetapir patterns across groups **(Figure 1, eTable 1** in the Supplement). Although the CSF-/PET+ group had more tracer uptake at baseline than CSF-/PET-, they did not accumulate significantly more amyloid-β over time on PET. Over time, CSF+/PET-group had widespread increase of tracer uptake compared to CSF-/PET-irrespective of the reference region. Additionally, CSF+/PET-had slightly more tracer uptake at baseline than CSF-/PET-.

**Figure 1.**
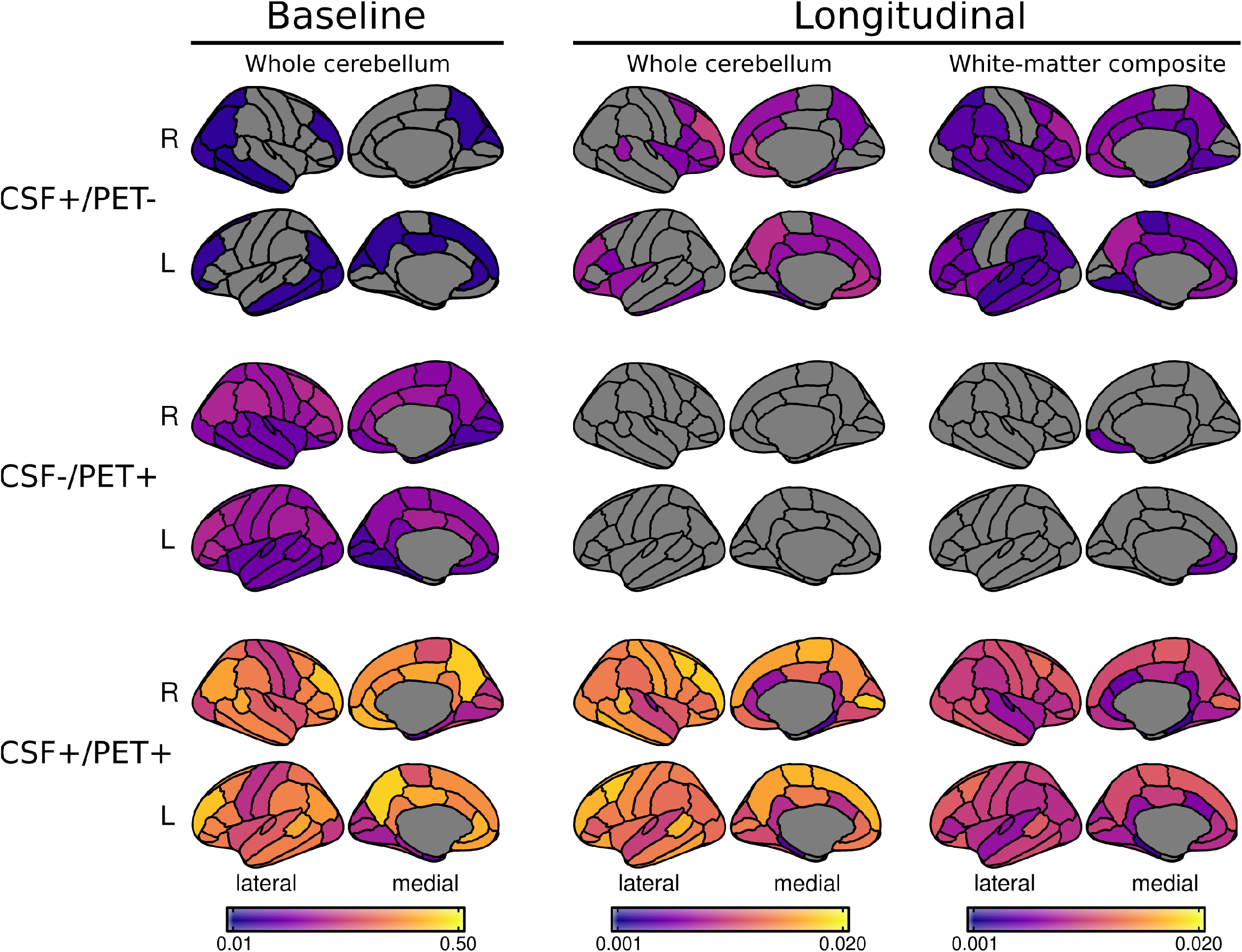
Accumulation of amyloid-β measured by [^18^F]florbetapir positron emission tomography. Results obtained from linear mixed models, with the colors indicating β-coefficients relative to the CSF-/PET-group. The three sections show the difference between the group of interest (CSF+/PET-, CSF-/PET+ or CSF+/PET+) and CSF-/PET-. The first column shows the β-coefficient for the baseline effect of the CSF/PET group when using whole cerebellum as the reference region. The second and third columns show the β-coefficients for the interaction between CSF/PET group and time as the longitudinal change when using whole cerebellum or composite white matter as the reference, respectively. Only regions with p<0.05 are shown. Image was created using the ggseg package in R.

### Longitudinal trajectories of tau and cognition

Next, we investigated longitudinal trajectories of tau pathology (CSF and PET) and cognition **(Figure 2, eTable 2** in the Supplement). Compared to amyloid-β CSF+/PET+ group, participants with discordant CSF/PET amyloid-β status had at baseline significantly (i) lower levels of CSF t-tau and p-tau measures (both p<0.001), (ii) less [^18^F]flortaucipir uptake in entorhinal cortex (both p<0.001), temporal meta-ROI (p<0.001 for CSF+/PET-, p=0.006 for CSF-/PET+) and Braak V/VI (p=0.002 for CSF+/PET-, p=0.274 for CSF-/PET+), and (iii) better cognitive test scores (MMSE: p=0.001 for CSF+/PET-, p=0.201 for CSF-/PET+; memory: both p<0.001; executive functioning: p=0.010 for CSF+/PET-, p<0.001 for CSF-/PET+). Longitudinally, participants in both CSF/PET discordant groups had lower rates of increase in [^18^F]flortaucipir uptake in temporal meta-ROI (p=0.010 for CSF+/PET-, p=0.031 for CSF-/PET+) and Braak V/VI composite areas (p=0.022 for CSF+/PET-), and less decline in cognitive test scores (all p<0.001 for both), compared to the CSF+/PET+ group. CSF+/PET-participants additionally had at baseline worse executive functioning than participants in the CSF-/PET-group (p=0.034). group. The CSF/PET discordant groups were otherwise similar to CSF-/PET-.

**Figure 2.**
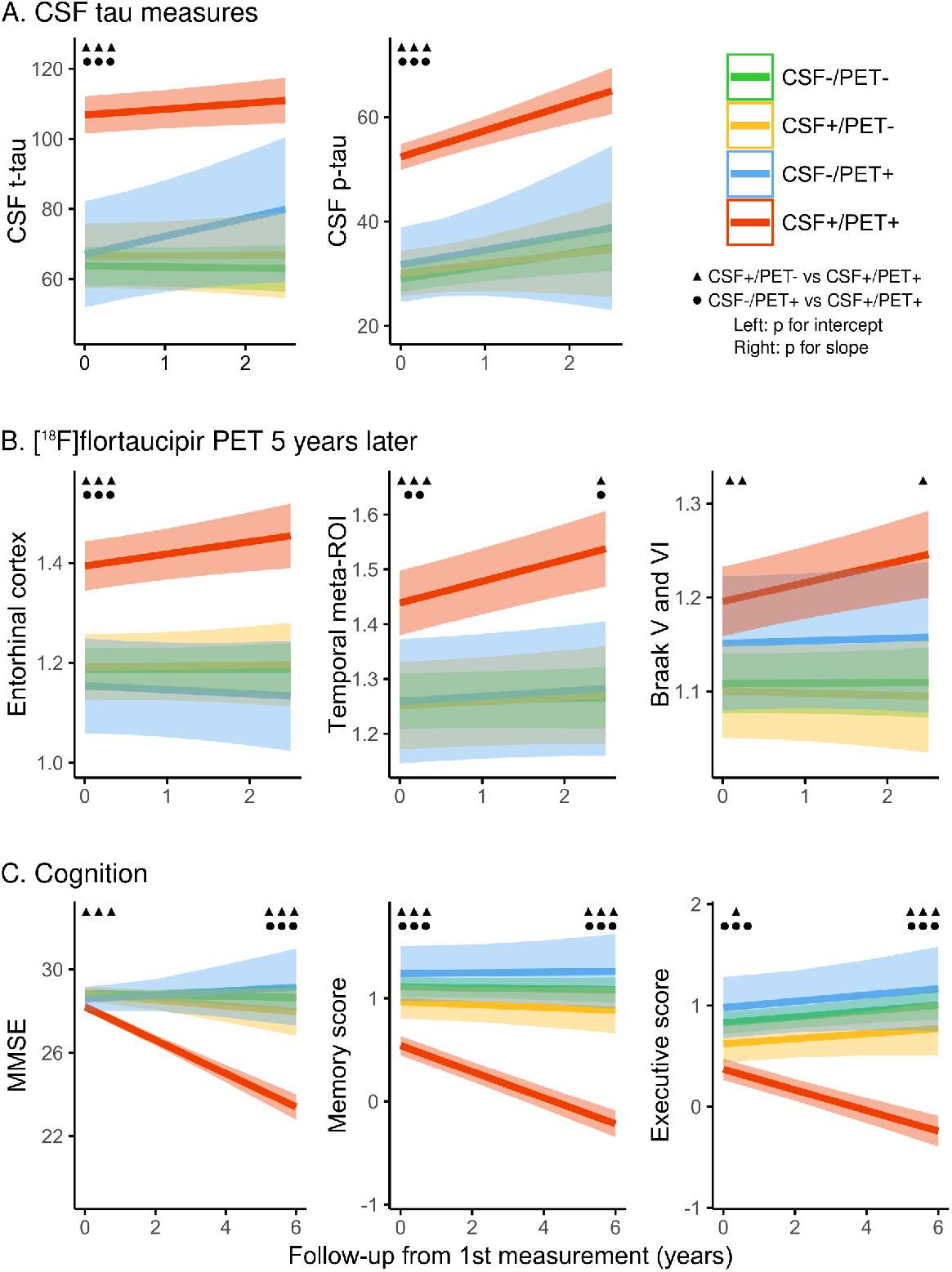
Longitudinal trajectories of tau pathology and cognition. Results obtained from linear mixed models investigating the effect of amyloid-β CSF/PET discordance on **A** tau pathology assessed by CSF t-tau and p-tau, **B** tau pathology based on regional [^18^F]flortaucipir PET, and **C** cognitive trajectories measured by MMSE, ADNI memory and executive composite scores. CSF tau was assessed from baseline to median 2.0 years; [^18^F]flortaucipir PET was first performed a median of 5.2 years after baseline and was followed up a median of 1.3 years later; cognitive tests were assessed at baseline, and followed up for a median of 4.2 years. Difference from CSF+/PET+ is illustrated by black triangles (for CSF+/PET-) or circles (for CSF-/PET+) with the number of symbols indicating statistical difference (one: p<0.05; two: p<0.01; three: p<0.001). Symbols on the left side of a plot show difference in intercept (main effect of CSF/PET group in the model), and symbols on the right side show a difference in slope (interaction between CSF/PET group and time). CSF+/PET-participants also had at baseline worse executive functioning than participants in the CSF-/PET-group (p=0.034). Image was created using the ggeffects package in R.

### Clinical Progression

Next, we investigated the association between amyloid-β CSF/PET status and clinical progression using Kaplan-Meier estimates and Cox regression analyses **(Figure 3, eTable 3** in the Supplement). Progression to either MCI or dementia occurred less often in amyloid-β CSF+/PET-(14%, hazard ratio [HR]= 0.22 [0.12, 0.41], p<0.001), CSF-/PET+ (8%, HR= 0.10 [0.02, 0.41], p=0.001) and CSF-/PET-(10%, HR= 0.16 [0.11, 0.24], p<0.001) participants, compared to the CSF+/PET+ (44%) group. Similarly, progression to dementia was rare in CSF+/PET-(5%, HR= 0.09 [0.03, 0.25], p<0.001) and CSF-/PET-(6%, HR= 0.11 [0.06, 0.17], p<0.001), but occurred more frequently in CSF+/PET+ (38%). No participants with amyloid-β CSF-/PET+ progressed to dementia.

**Figure 3.**
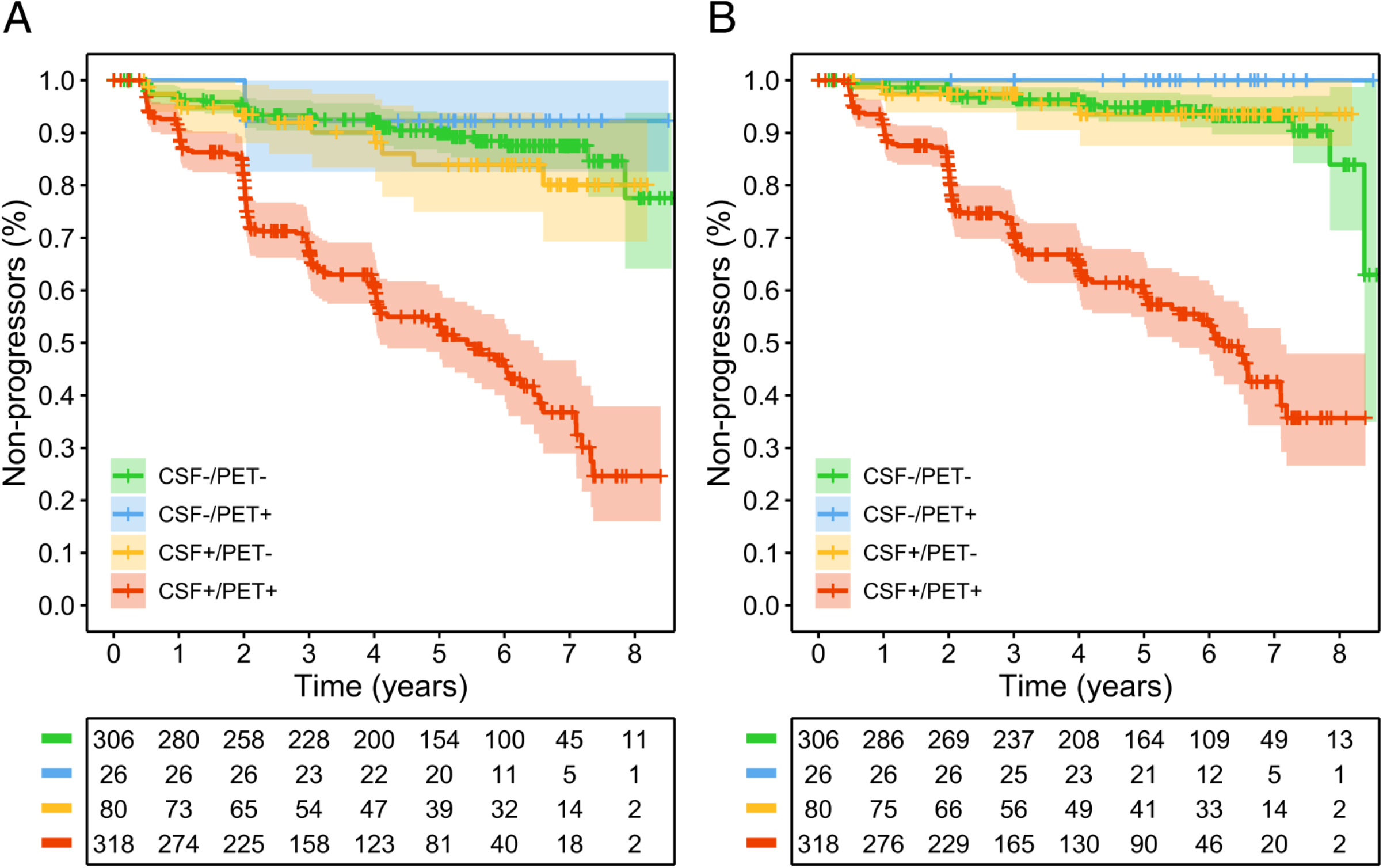
Kaplan-Meier curves for clinical progression. Results obtained from the Kaplan-Meier estimate investigating the association between amyloid-β CSF/PET profile and clinical progression from CN to MCI or dementia, or from MCI to dementia (A) and progression from CN or MCI to dementia (B). Tables below the figures report per year the number of participants with available follow-up data.

### Associations between amyloid-β and tau in CSF+/PET-participants

We then assessed whether continuous [^18^F]florbetapir uptake in the subthreshold range is correlated with tau measures in the amyloid-β CSF+/PET-group **(Figure 4)**. [^18^F]florbetapir tracer uptake globally and in early accumulating regions was associated with higher baseline CSF t-tau (p=0.001 and p=0.003), and higher [^18^F]flortaucipir uptake in entorhinal cortex (p=0.003 and p=0.010) and marginally in the temporal meta-ROI (p=0.062 and p=0.091).

**Figure 4.**
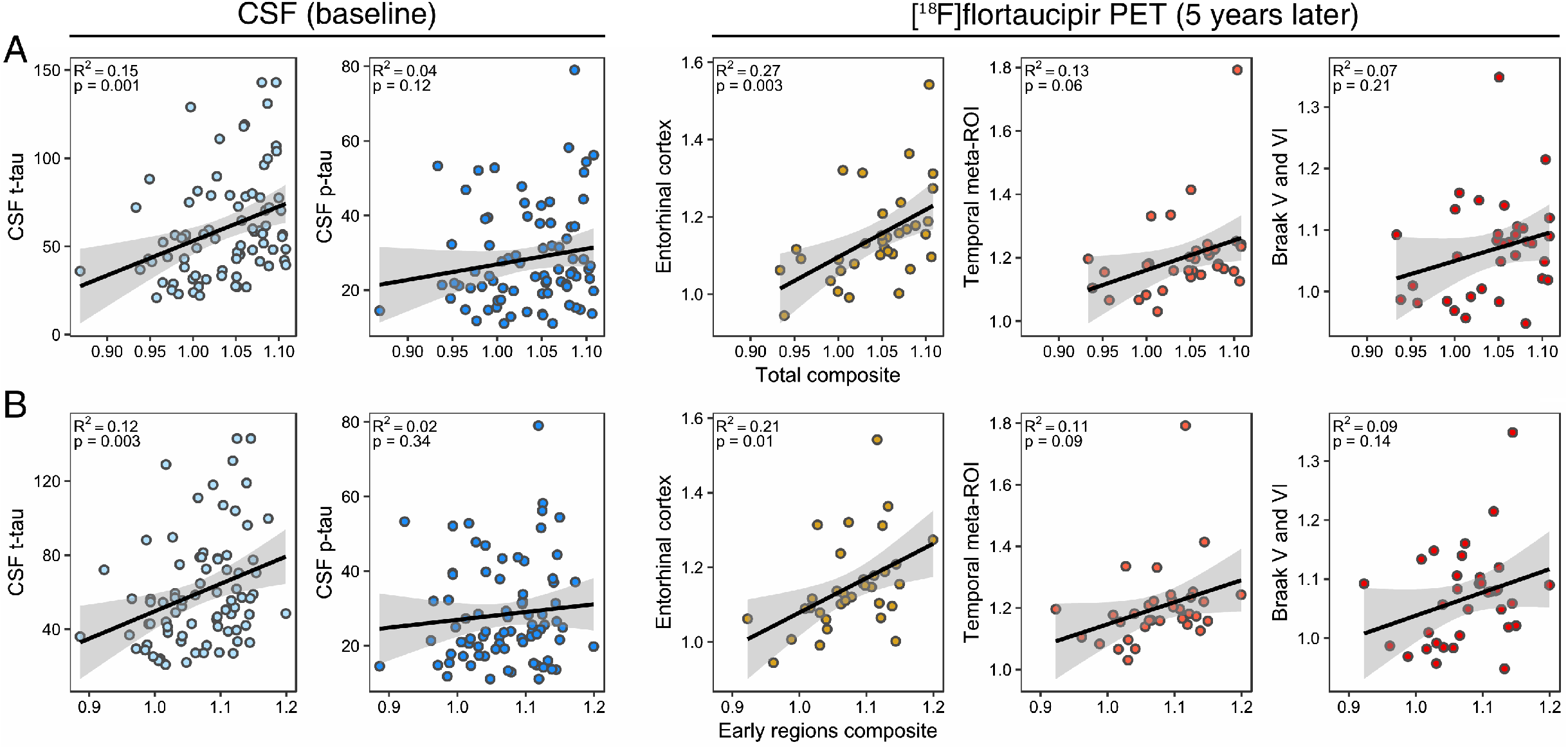
Correlation between baseline [^18^F]florbetapir PET and tau pathology based on CSF and PET. Plotted is the association between [^18^F]florbetapir PET, based on **A** total composite and **B** early accumulating regions composite on the x axis, and cross-sectional tau pathology, measured by either CSF t-tau or p-tau at baseline or [^18^F]flortaucipir PET median of 5.2 years after baseline on the y axis. R^2^ and p values are reported from the from linear regression models, which were also adjusted for the times between baseline [^18^F]florbetapir PET and the outcome modality.

### Accumulation of amyloid-β and tau in CSF+/PET-participants

Finally, we investigated whether CSF+/PET-participants progress to CSF+/PET+ or become tau-positive first. Based on the last available [^18^F]florbetapir amyloid-β PET scan and CSF Aβ42 analysis, 21/64 (33%) of the baseline amyloid-β CSF+/PET-participants progressed to CSF+/PET+. Of 34 CSF+/PET-participants with [^18^F]flortaucipir PET available, 11/34 (32% [95% confidence interval: 17%, 48%]) progressed to CSF+/PET+, but only one (3% [95% confidence interval: -3%, 9%], difference in proportions p<0.05) turned tau-positive based on entorhinal or temporal meta-ROI regions, and none in Braak V/VI. For comparison, 2/123 (2%, p=1.00) of amyloid-β CSF-/PET-, 0/16 (0%, p=1.00) CSF-/PET+, and 47/80 CSF+/PET+ (59%, p<0.001) were tau-positive based on last available [^18^F]flortaucipir PET.

## Discussion

We investigated the association between discordant CSF/PET amyloid-β biomarkers on tau pathology and clinical progression. Our main finding was that although amyloid-β CSF+/PET-participants showed longitudinal accumulation of amyloid-β based on PET, they had significantly less tau pathology based on both CSF at baseline and [^18^F]flortaucipir 5 years later compared to participants with CSF+/PET+ amyloid-β status. Similarly, discordant amyloid-β status was associated with better cognitive outcome and a lower risk of clinical progression than CSF+/PET+. We also showed that during follow-up, CSF+/PET-subjects frequently progressed to amyloid-β CSF+/PET+, whereas only one participant reached the threshold of tau-positivity based on [^18^F]flortaucipir PET. Finally, we showed a correlation between tau measures and global amyloid-β PET tracer uptake, indicating possible subthreshold accumulation of AD pathology in CSF+/PET-subjects. Taken together, our findings suggest that CSF+/PET-amyloid-β status is associated with a distinctly better prognosis than CSF+/PET+, and that a sufficient amyloid-β load detectable by both CSF and PET is required before significant tau deposition emerges.

Using both CSF tau measures and [^18^F]flortaucipir PET we found that participants with discordant CSF/PET amyloid-β status had less tau pathology than CSF+/PET+ amyloid-β, and comparable tau load as observed in concordant amyloid-β negative participants. It has been proposed that CSF+/PET-status can be caused by CSF Aβ42 being able to detect amyloid-β at an earlier stage due to the decrease of soluble Aβ42 in CSF preceding fibrillary depositions visualized by PET. This is supported by higher rates of CSF+/PET-compared to CSF-/PET+ across several studies.^7,37,38^ Previous longitudinal PET studies have shown that subjects with CSF+/PET-amyloid-β status show significant accumulation of amyloid-β over time.^8,9,39^ We replicated this finding in our study, further supporting the notion that CSF+/PET-amyloid-β status identifies the beginnings of amyloid-β accumulation. Although participants with CSF-/PET+ amyloid-β status had higher [^18^F]florbetapir tracer uptake at baseline, they did not show significant accumulation of amyloid-β over time. Combined with the lack of clinical progression in this group, these observations suggest that isolated amyloid-β PET positivity might be caused by non-specific tracer uptake in the white matter, processing errors or other unknown factors.^6^ In CSF+/PET-participants, the lack of substantial tau pathology based on CSF tau measures at baseline and on [^18^F]flortaucipir PET five years later, accompanied by lack of cognitive decline and clinical progression, suggests these subjects have a distinctly more favorable prognosis than subjects with CSF+/PET+. This is likely caused by the remarkably slow course of AD, which is characterized by gradual accumulation of pathology over time.^40,41^ Accounting for the more benign prognosis of CSF+/PET-subjects is important for the timing of future interventions at the earliest stages of AD pathology.

Current hypothetical biomarker models suggest that accumulation of amyloid-β pathology is followed by detectable cortical tau pathology, subsequently leading to neurodegeneration and cognitive decline.^42,43^ Although it has been proposed that CSF+/PET-is followed by conversion to CSF+/PET+,^8,44^ the exact timing of CSF+/PET-status in regard to that timeline, in particular towards accumulation of tau, is unknown. We found that within five years, one third of the CSF+/PET-participants progressed to CSF+/PET+, whereas at that time only one participant exhibited suprathreshold early to intermediate stage tau pathology based on [^18^F]flortaucipir PET and none showed widespread neocortical uptake (i.e. Braak stage V/VI regions). This finding has at least two implications. First, as the majority of CSF+/PET-participants did not progress to CSF+/PET+ within 5 years, this indicates that in the majority of cases the CSF+/PET-amyloid-β status lasts for several years. Second, accumulation of sufficient amyloid-β detectable by both CSF and PET seems to precede significant accumulation of tau pathology.^45^ However, we also found a correlation between baseline regional amyloid-β PET and tau pathology in the CSF+/PET-group, suggesting that there already might be interaction present between amyloid-β and tau. This supports previous work, emphasizing the importance of considering subthreshold accumulation of pathology to better understand disease mechanisms of early preclinical stages of AD.^46–48^

Our study has some limitations. Although ADNI is one of the largest cohorts with both available amyloid-β PET and CSF analysis, only a relatively small number of participants with discordant CSF/PET amyloid-β status were available. Second, our main outcome measures of tau pathology based on CSF and PET were assessed at different time-points. Although that reduces the direct comparability of these findings, they also complement each other and allow to measure tau pathology both at baseline and several years later. Additionally, relatively short follow-up periods were available for both CSF tau measures and [^18^F]flortaucipir PET. Therefore, it is possible, that with longer follow-up periods, subjects with discordant amyloid-β status might show diverging trajectories compared to the CSF-/PET-group. Our interpretation of the study could also be afflicted by the possibility that CSF+/PET-status might reflect a different subtype of AD, although no evidence for that exists. Finally, cut-offs of biomarkers as well as defining amyloid-β PET status based on global SUVR are important considerations when evaluating these results. As suboptimal cut-offs might result in misclassification,^49^ we used applied widely used and validated cut-offs for both PET and CSF.

In conclusion, our findings indicate that a sufficient amyloid-β load detectable by both PET and CSF is required before substantial tau deposition emerges. Subjects with CSF+/PET-amyloid-β profile are at a significantly earlier clinical and biological disease stage than those with CSF+/PET+, and have a distinctly better prognosis for at least five years.

## Acknowledgements

Data collection and sharing for this project was funded by the Alzheimer’s Disease Neuroimaging Initiative (ADNI) (National Institutes of Health Grant U01 AG024904) and DOD ADNI (Department of Defense award number W81XWH-12-2-0012). ADNI is funded by the National Institute on Aging, the National Institute of Biomedical Imaging and Bioengineering, and through generous contributions from the following: AbbVie, Alzheimer’s Association; Alzheimer’s Drug Discovery Foundation; Araclon Biotech; BioClinica, Inc.; Biogen; Bristol-Myers Squibb Company; CereSpir, Inc.; Cogstate; Eisai Inc.; Elan Pharmaceuticals, Inc.; Eli Lilly and Company; EuroImmun; F. Hoffmann-La Roche Ltd and its affiliated company Genentech, Inc.; Fujirebio; GE Healthcare; IXICO Ltd.; Janssen Alzheimer Immunotherapy Research & Development, LLC.; Johnson & Johnson Pharmaceutical Research & Development LLC.; Lumosity; Lundbeck; Merck & Co., Inc.; Meso Scale Diagnostics, LLC.; NeuroRx Research; Neurotrack Technologies; Novartis Pharmaceuticals Corporation; Pfizer Inc.; Piramal Imaging; Servier; Takeda Pharmaceutical Company; and Transition Therapeutics. The Canadian Institutes of Health Research is providing funds to support ADNI clinical sites in Canada. Private sector contributions are facilitated by the Foundation for the National Institutes of Health (www.fnih.org). The grantee organization is the Northern California Institute for Research and Education, and the study is coordinated by the Alzheimer’s Therapeutic Research Institute at the University of Southern California. ADNI data are disseminated by the Laboratory for Neuro Imaging at the University of Southern California.

The Alzheimer Center Amsterdam is supported by Alzheimer Nederland and Stichting VUmc fonds. Research performed at the Alzheimer Center Amsterdam is part of the neurodegeneration research program of Amsterdam Neuroscience. J.R. would like to thank Sergei Nazarenko, the International Atomic Energy Agency (IAEA) and the North Estonia Medical Centre for their contribution to his professional development. The authors would like to acknowledge Nicholas Cullen for his support in statistical analysis.

P.S. received grants from GE Healthcare, Piramal, and Merck, paid to his institution; he has received speaker’s fees paid to the institution Alzheimer Center, VU University Medical Center, Lilly, GE Healthcare, and Roche. The funding sources were not involved in the writing of this article or in the decision to submit it for publication. J.R., L.C., F.B., and R.O. report no competing interests.

## References

1. Braak H, Braak E. Neuropathological stageing of Alzheimer-related changes. Acta Neuropathol. 1991;82:239–259. doi:10.1007/BF00308809

2. Jack CR, Bennett DA, Blennow K, et al. NIA-AA Research Framework: Toward a biological definition of Alzheimer’s disease. Alzheimer’s Dement. 2018;14(4):535–562. doi:10.1016/j.jalz.2018.02.018

3. Albert MS, DeKosky ST, Dickson D, et al. The diagnosis of mild cognitive impairment due to Alzheimer’s disease: Recommendations from the National Institute on Aging-Alzheimer’s Association workgroups on diagnostic guidelines for Alzheimer’s disease. Alzheimer’s Dement. 2011;7(3):270–279. doi:10.1016/j.jalz.2011.03.008

4. McKhann GM, Knopman DS, Chertkow H, et al. The diagnosis of dementia due to Alzheimer’s disease: Recommendations from the National Institute on Aging-Alzheimer’s Association workgroups on diagnostic guidelines for Alzheimer’s disease. Alzheimer’s Dement. 2011;7(3):263–269. doi:10.1016/j.jalz.2011.03.005

5. Zwan M, van Harten A, Ossenkoppele R, et al. Concordance between cerebrospinal fluid biomarkers and [11C]PIB PET in a memory clinic cohort. J Alzheimers Dis. 2014;41(3):801–807. doi:10.3233/JAD-132561

6. Mattsson N, Insel PS, Donohue M, et al. Independent information from cerebrospinal fluid amyloid-β and florbetapir imaging in Alzheimer’s disease. Brain. 2015;138(3):772–783. doi:10.1093/brain/awu367

7. de Wilde A, Reimand J, Teunissen CE, et al. Discordant amyloid-β PET and CSF biomarkers and its clinical consequences. Alzheimers Res Ther. 2019;11(1):78. doi:10.1186/s13195-019-0532-x

8. Palmqvist S, Mattsson N, Hansson O. Cerebrospinal fluid analysis detects cerebral amyloid-β accumulation earlier than positron emission tomography. Brain. 2016;139(4):1226–1236. doi:10.1093/brain/aww015

9. Palmqvist S, Schöll M, Strandberg O, et al. Earliest accumulation of β-amyloid occurs within the default-mode network and concurrently affects brain connectivity. Nat Commun. 2017;8(1). doi:10.1038/s41467-017-01150-x

10. Blennow K, Wallin A, Ågren H, Spenger C, Siegfried J, Vanmechelen E. tau protein in cerebrospinal fluid. Mol Chem Neuropathol. 1995;26(3):231–245. doi:10.1007/BF02815140

11. Chien DT, Bahri S, Szardenings AK, et al. Early Clinical PET Imaging Results with the Novel PHF-Tau Radioligand [F-18]-T807. J Alzheimer’s Dis. 2013;34(2):457–468. doi:10.3233/JAD-122059

12. Lowe VJ, Wiste HJ, Senjem ML, et al. Widespread brain tau and its association with ageing, Braak stage and Alzheimer’s dementia. Brain. 2018;141(1):271–287. doi:10.1093/brain/awx320

13. Ossenkoppele R, Schonhaut DR, Schöll M, et al. Tau PET patterns mirror clinical and neuroanatomical variability in Alzheimer’s disease. Brain. 2016;139(5):1551–1567. doi:10.1093/brain/aww027

14. Bejanin A, Schonhaut DR, La Joie R, et al. Tau pathology and neurodegeneration contribute to cognitive impairment in Alzheimer’s disease. Brain. 2017;140(12):3286–3300. doi:10.1093/brain/awx243

15. Hardy J, Bogdanovic N, Winblad B, et al. Pathways to Alzheimer’s disease. J Intern Med. 2014;275(3):296–303. doi:10.1111/joim.12192

16. Petersen RC, Aisen PS, Beckett LA, et al. Alzheimer’s Disease Neuroimaging Initiative (ADNI): Clinical characterization. Neurology. 2010;74(3):201–209. doi:10.1212/WNL.0b013e3181cb3e25

17. Crane PK, Carle A, Gibbons LE, et al. Development and assessment of a composite score for memory in the Alzheimer’s Disease Neuroimaging Initiative (ADNI). Brain Imaging Behav. 2012;6(4):502–516. doi:10.1007/s11682-012-9186-z

18. Gibbons LE, Carle AC, Mackin RS, et al. A composite score for executive functioning, validated in Alzheimer’s Disease Neuroimaging Initiative (ADNI) participants with baseline mild cognitive impairment. Brain Imaging Behav. 2012;6(4):517–527. doi:10.1007/s11682-012-9176-1

19. Landau S, Jagust W. Florbetapir processing methods. ADNI. 2011.

20. Jagust WJ, Landau SM, Koeppe RA, et al. The Alzheimer’s Disease Neuroimaging Initiative 2 PET Core: 2015. Alzheimer’s Dement. 2015;11(7):757–771. doi:10.1016/j.jalz.2015.05.001

21. Landau SM, Breault C, Joshi AD, et al. Amyloid-Imaging with Pittsburgh Compound B and Florbetapir: Comparing Radiotracers and Quantification Methods. J Nucl Med. 2013;54(1):70–77. doi:10.2967/jnumed.112.109009

22. Clark CM, Pontecorvo MJ, Beach TG, et al. Cerebral PET with florbetapir compared with neuropathology at autopsy for detection of neuritic amyloid-β plaques: A prospective cohort study. Lancet Neurol. 2012;11(8):669–678. doi:10.1016/S1474-4422(12)70142-4

23. Joshi AD, Pontecorvo MJ, Clark CM, et al. Performance characteristics of amyloid PET with florbetapir F 18 in patients with Alzheimer’s disease and cognitively normal subjects. J Nucl Med. 2012;53(3):378–384. doi:10.2967/jnumed.111.090340

24. Landau SM, Fero A, Baker SL, et al. Measurement of Longitudinal -Amyloid Change with 18F-Florbetapir PET and Standardized Uptake Value Ratios. J Nucl Med. 2015;56(4):567–574. doi:10.2967/jnumed.114.148981

25. Mattsson N, Palmqvist S, Stomrud E, Vogel J, Hansson O. Staging β -Amyloid Pathology With Amyloid Positron Emission Tomography. JAMA Neurol. 2019. doi:10.1001/jamaneurol.2019.2214

26. Shaw LM, Vanderstichele H, Knapik-Czajka M, et al. Cerebrospinal fluid biomarker signature in alzheimer’s disease neuroimaging initiative subjects. Ann Neurol. 2009;65(4):403–413. doi:10.1002/ana.21610

27. Shaw LM, Vanderstichele H, Knapik-Czajka M, et al. Qualification of the analytical and clinical performance of CSF biomarker analyses in ADNI. Acta Neuropathol. 2011;121(5):597–609. doi:10.1007/s00401-011-0808-0

28. Landau S, Jagust W. Flortaucipir (AV-1451) processing methods. ADNI. 2018.

29. Jack CR, Wiste HJ, Weigand SD, et al. Defining imaging biomarker cut points for brain aging and Alzheimer’s disease. Alzheimer’s Dement. 2017;13(3):205–216. doi:10.1016/j.jalz.2016.08.005

30. Schöll M, Lockhart SN, Schonhaut DR, et al. PET Imaging of Tau Deposition in the Aging Human Brain. Neuron. 2016;89(5):971–982. doi:10.1016/j.neuron.2016.01.028

31. Johnson KA, Schultz A, Betensky RA, et al. Tau positron emission tomographic imaging in aging and early Alzheimer disease. Ann Neurol. 2016;79(1):110–119. doi:10.1002/ana.24546

32. Ossenkoppele R, Palmqvist S, Mattsson N, Janelidze S, Santillo A, Ohlsson T. Discriminative Accuracy of [18F]flortaucipir Positron Emission Tomography for Alzheimer Disease vs Other Neurodegenerative Disorders. 2018. doi:10.1001/jama.2018.12917

33. R Core Team. R: A language and environment for statistical computing. 2018:https://www.R-project.org/.

34. Bates D, Mächler M, Bolker BM, Walker SC. Fitting linear mixed-effects models using lme4. J Stat Softw. 2015. doi:10.18637/jss.v067.i01

35. Mowinckel A, Pineiro DV. ggseg package for R. 2018. https://github.com/LCBC-UiO/ggseg.

36. Lüdecke D. ggeffects: Tidy Data Frames of Marginal Effects from Regression Models. J Open Source Softw. 2018. doi:10.21105/joss.00772

37. Fagan AM, Mintun MA, Shah AR, et al. Cerebrospinal fluid tau and ptau181 increase with cortical amyloid deposition in cognitively normal individuals: Implications for future clinical trials of Alzheimer’s disease. EMBO Mol Med. 2009;1(8-9):371–380. doi:10.1002/emmm.200900048

38. Blennow K, Mattsson N, Schöll M, Hansson O, Zetterberg H. Amyloid biomarkers in Alzheimer’s disease. Trends Pharmacol Sci. 2015;36(5):297–309. doi:10.1016/j.tips.2015.03.002 Review

39. Vlassenko AG, McCue L, Jasielec MS, et al. Imaging and cerebrospinal fluid biomarkers in early preclinical alzheimer disease. Ann Neurol. 2016;80(3):379–387. doi:10.1002/ana.24719

40. Villemagne VL, Burnham S, Bourgeat P, et al. Amyloid β deposition, neurodegeneration, and cognitive decline in sporadic Alzheimer’s disease: A prospective cohort study. Lancet Neurol. 2013;12(4):357–367. doi:10.1016/S1474-4422(13)70044-9

41. Jack CR, Wiste HJ, Schwarz CG, et al. Longitudinal tau PET in ageing and Alzheimer’s disease. Brain. 2018;141(5):1517–1528. doi:10.1093/brain/awy059

42. Jack C. Hypothetical Pathological Cascade in Alheimer’s Disease. Lancet, The. 2010;9(1):1–20. doi:10.1016/S1474-4422(09)70299-6.Hypothetical

43. Jack CR, Holtzman DM. Biomarker modeling of alzheimer’s disease. Neuron. 2013;80(6):1347–1358. doi:10.1016/j.neuron.2013.12.003

44. Reimand J, de Wilde A, Teunissen CE, et al. PET and CSF amyloid-β status are differently predicted by patient features: information from discordant cases. Alzheimers Res Ther. 2019;11(1):100. doi:10.1186/s13195-019-0561-5

45. Jack CR, Wiste HJ, Botha H, et al. The bivariate distribution of amyloid-β and tau: relationship with established neurocognitive clinical syndromes. Brain. 2019;142(10):3230–3242. doi:10.1093/brain/awz268

46. Leal SL, Lockhart SN, Maass A, Bell RK, Jagust WJ. Subthreshold amyloid predicts tau deposition in aging. J Neurosci. 2018;38(19):4482–4489. doi:10.1523/JNEUROSCI.0485-18.2018

47. Landau SM, Horng A, Jagust WJ. Memory decline accompanies subthreshold amyloid accumulation. Neurology. 2018;90(17):E1452–E1460. doi:10.1212/WNL.0000000000005354

48. Insel PS, Ossenkoppele R, Gessert D, et al. Time to Amyloid Positivity and Preclinical Changes in Brain Metabolism, Atrophy, and Cognition: Evidence for Emerging Amyloid Pathology in Alzheimer’s Disease. Front Neurosci. 2017;11(MAY):1–9. doi:10.3389/fnins.2017.00281

49. McRae-McKee K, Udeh-Momoh CT, Price G, et al. Perspective: Clinical relevance of the dichotomous classification of Alzheimer’s disease biomarkers: Should there be a “gray zone”? Alzheimer’s Dement. 2019;15(10):1348–1356. doi:10.1016/j.jalz.2019.07.010

